# BUDAPEST: A Fast and Reliable Bayesian Algorithm for TMS Threshold Estimation with an Open-Source GUI and Human Validation

**DOI:** 10.64898/2026.03.03.26347528

**Authors:** Danyal Fareed Bhutto, Evgenii Kim, Netri Pajankar, Farzan Vahedifard, Mohammad Daneshzand, Dylan Edwards, Aapo Nummenmaa

**Affiliations:** Athinoula A. Martinos Center for Biomedical Imaging, Department of Radiology, Massachusetts General Hospital, Boston, MA, USA; Harvard Medical School, Boston, MA, USA; Jefferson Moss Rehabilitation Research Institute, Philadelphia, PA, USA; Thomas Jefferson University, Department of Rehabilitation Medicine, Philadelphia, PA, USA

**Keywords:** Transcranial magnetic stimulation, Neuronavigation, Dosing, Parameter Estimation

## Abstract

**Background:** Motor threshold (MT) estimation is fundamental to transcranial magnetic stimulation (TMS), guiding individualized stimulation intensity in research and therapy. Conventional methods such as the 5-out-of-10 rule require many stimuli, while adaptive approaches like Parameter Estimation by Sequential Testing (PEST) improve efficiency but can exhibit poor convergence under certain conditions.

**Objective:** This study introduces the Bayesian Uncertainty Dynamic Algorithm for Parameter Estimation by Sequential Testing (BUDAPEST), a Bayesian adaptive method for fast, accurate MT estimation with user-controlled uncertainty. The aims were to validate its accuracy in simulations and human data, promote usability through a MATLAB-based graphical interface, and evaluate experimental utility through resting and active MT comparisons and session-to-session reliability.

**Methods:** BUDAPEST infers MT from binary MEP responses using sequential Bayesian updating and terminates when a user-defined uncertainty threshold is reached. Performance was evaluated in 10,000 virtual simulations and in human rMT and aMT measurements across two sessions per subject, including 3×5 cortical motor mapping to assess physiological spatial patterns.

**Results:** In simulations, BUDAPEST achieved a mean absolute error of 1.9% MSO within ~10 pulses using a 2% uncertainty criterion while avoiding PEST misestimations. In human data, MT estimates were accurate within ±4% MSO and robust to initialization; rMT showed strong session-to-session reliability (r = 0.78), whereas aMT exhibited greater variability. Motor mapping revealed coherent excitability gradients centered on the hotspot.

**Conclusion:** BUDAPEST enables rapid, reliable, and uncertainty-controlled MT estimation while reducing procedure time and participant burden. The accompanying GUI facilitates immediate adoption in research and clinical TMS environments.

**Highlights:**

- Introduces BUDAPEST, a Bayesian uncertainty-aware algorithm for rapid and reliable TMS motor threshold estimation.
- Achieves accurate MT estimates (≈2% MSO error) in ~10 pulses with user-controlled trade-offs between precision and procedure duration.
- Demonstrates robust performance in simulations and human data, with strong resting MT reliability and an open-source GUI enabling immediate adoption.

## Introduction

Transcranial Magnetic Stimulation (TMS) is a noninvasive technique commonly used to map the human motor cortex. It modulates neuronal activity by generating time-varying magnetic fields that induce electric fields (E-fields) in targeted brain regions [1,2]. Owing to its ability to safely and painlessly induce suprathreshold E-fields within the intracranial space, TMS has been widely adopted in research and clinical contexts [3,4], including FDA-approved treatments for disorders such as treatment-resistant depression [5] and obsessive-compulsive disorder (OCD) [6], as well as presurgical functional mapping of motor and language regions [7]. In the context of hardware development, optimization of TMS coil design remains an active area of research, as tailoring coil geometries for specific applications directly influences stimulation intensity requirements and contributes to variability across studies [8,9]. In parallel, multichannel transcranial magnetic stimulation (mTMS) arrays have emerged as a promising approach, enabling multiple cortical sites to be stimulated simultaneously or sequentially under electronic control without mechanical repositioning of the stimulation coils [10].

Accurate and reliable identification of the optimal TMS site and stimulation intensity is critical for all applications. In motor cortex studies, TMS applied above a certain intensity elicits a measurable electrical response in the muscle contralateral to the stimulated cortical neurons, termed the motor-evoked potential (MEP). The International Federation of Clinical Neurophysiology (IFCN) defines the motor threshold (MT) intensity as the minimum TMS output intensity required to evoke reliable MEPs (typically in resting state ≥50 µV) in 50% of trials [11]. Even though the excitability of different cortical areas is expected to vary, the resting motor threshold (rMT) acts as standard reference for setting optimal stimulation parameters.

Several protocols exist for MT assessment; one widely cited by the IFCN is the “5-out-of-10” method. In this protocol, stimulation begins at either a subthreshold or suprathreshold intensity, and the experimenter records the presence or absence of a motor response. If a response occurs in more than five of ten trials, the intensity is decreased by 5%; if fewer, the intensity is increased by 5%. Once the response is observed in at least five out of ten trials, the intensity is adjusted in 2% increments, followed by 1% increments, until the lowest intensity producing responses in five out of ten trials is identified. This final intensity defines the MT and the corresponding percentage of maximum stimulator output (%MSO) [12]. This systematic protocol typically requires a large number of stimuli, averaging approximately 50 to 75 TMS pulses to determine the MT accurately [13–15].

Reducing the duration of MT determination has the potential to decrease experimental time, minimize participant discomfort, and limit physiological changes associated with repeated TMS pulses, prolonged stimulation, or repeated muscle activation during active MT (aMT) measurements, including increases in motor map area and volume [16]. To reduce the number of stimulation trials, adaptive methods based on Parameter Estimation by Sequential Testing (PEST) combined with maximum likelihood regression have been developed [17], enabling MT estimation with approximately 14–16 pulses at a 95% confidence interval and up to 30 pulses for full confidence, while maintaining accuracy within 5% of MSO [14,18].

A major limitation of the traditional PEST algorithm is its susceptibility to misestimation, particularly severe underestimation, without adequate correction even when subsequent MEP responses contradict earlier predictions. For example, if an early MEP is elicited at an intensity below the true MT, PEST may overweight this observation and continue sampling excessively low stimulator outputs, ignoring later evidence of absent MEPs. Although PEST is generally reported to achieve MT estimates within 5% of MSO, this behavior raises concerns. In a worst-case scenario, PEST was shown to underestimate the rMT by as much as 23% MSO after 14 pulses, and 16% MSO below the true value after 30 pulses (true MT = 71% MSO). Even when using the “refinement of a raw threshold estimate” mode, reported errors still range from –7% to +9% MSO [19].

Bayesian adaptations of PEST and machine learning–based approaches for MT estimation have been proposed, yet their adoption in experimental and clinical TMS settings remain limited [20,21]. An adaptive method currently under clinical investigation is the Stochastic Approximator of Motor Threshold (SAMT), which estimates MT using a model-free stochastic approximation procedure that updates stimulation intensity based on binary suprathreshold responses. SAMT adjusts pulse amplitude according to a predefined, decreasing step-size schedule and assesses convergence by achieving an approximately 50% response rate, with accuracy and confidence evaluated retrospectively by fitting a sigmoidal response model to the collected data. Reported performance indicates convergence in approximately 25 trials with an average error of ~1.3% MSO relative to the fitted threshold [22].

A key remaining challenge in MT determination is the lack of principled online stopping criteria that provide quantitative guidance during the estimation process. Although SAMT has demonstrated clinical feasibility and rapid convergence, it targets the 50% response level using predefined stochastic approximation step-size rules rather than explicitly quantifying uncertainty during the procedure. While SAMT adapts stimulation direction and step size based on observed responses, it does not provide trial-by-trial estimates of confidence or uncertainty in the threshold estimate [22]. This limitation is important, as different experimental and clinical applications impose distinct requirements on accuracy and procedure duration, underscoring the need for MT estimation frameworks that explicitly quantify uncertainty and allow users to tailor stopping criteria accordingly.

In this study, we introduce the Bayesian Uncertainty Dynamic Algorithm for Parameter Estimation by Sequential Testing (BUDAPEST), a principled Bayesian active-learning framework that models the MT as a statistical parameter subject to uncertainty, and sequentially updates its posterior distribution based on binary MEP responses to TMS pulses. This work has three primary objectives: (1) to establish the methodological foundation of BUDAPEST and quantify its accuracy using both virtual simulations and human data; (2) to promote practical adoption by integrating BUDAPEST into an intuitive MATLAB-based graphical user interface (GUI), that guides experimenters through the MT procedure using user-defined uncertainty thresholds (see Figure 1), enabling flexible control over the trade-off between estimation accuracy and procedure duration; and (3) to evaluate its experimental utility by comparing rMT and aMT measurements, characterizing session-to-session variability, and interpreting the physiological spatial mapping results. By directly modeling uncertainty and adapting sampling in real time, BUDAPEST overcomes long-standing limitations of traditional adaptive thresholding methods, improving reliability, reducing stimulation burden, and enabling reproducible MT estimation for both research and clinical applications.

**Figure 1.**
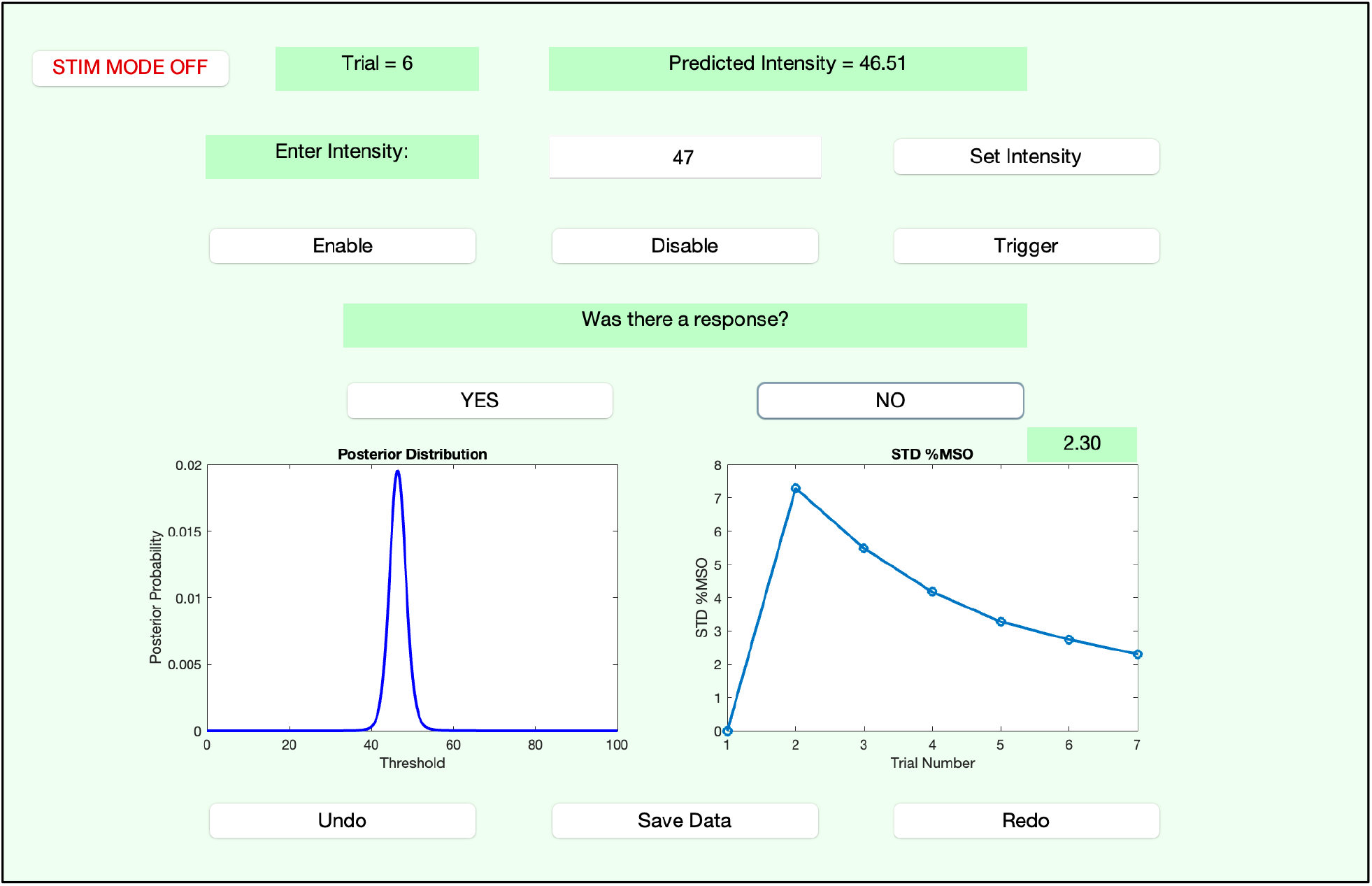
MATLAB-based GUI for the BUDAPEST Algorithm. The interface displays the current trial number and predicted intensity MT (%MSO), allows manual entry of measured MEP responses, and updates the posterior distribution (left panel) and uncertainty (right panel) in real time. The GUI provides visual guidance during threshold estimation and supports user-defined uncertainty-based stopping criteria. In addition, the GUI includes serial port communication capability, enabling direct control of MagVenture TMS stimulators for automated pulse delivery.

## Methods

### Bayesian Framework

The experimenter specifies an initial stimulation intensity and a stopping criterion defined as an acceptable level of posterior uncertainty (i.e., a user-defined standard deviation in %MSO). The algorithm initializes a prior probability distribution over candidate MT values using a Gaussian function centered at the starting intensity, with a scaling factor of *k*= 0.5 and a width spanning ±30% MSO to allow broad initial uncertainty. These parameters were empirically selected using preliminary virtual-subject simulations [19]. During the estimation process, the posterior distribution is iteratively updated after each trial according to Bayes’ rule based on the observed binary MEP response. The range of candidate MT values is dynamically adjusted as the posterior converges, refining the search space around the most probable MT estimate.

The subject’s response at each trial is binary with a response indicated as 1 and no response indicated as 0. These binary outcomes are assumed to arise from a sigmoid (logistic) function of the stimulation intensity relative to the true threshold. The sigmoid function provides a probabilistic mapping from intensity to response probability, with its inflection point (the 50% response probability) corresponding to the estimated MT:

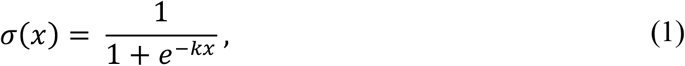

where *σ(x)*is the sigmoid function and *k*= 0.5is the steepness parameter of the sigmoid curve. The likelihood of observing *r* ∈ {0,1} binary response given threshold *θ* and intensity delivered *I* for a given trial can be modeled as:

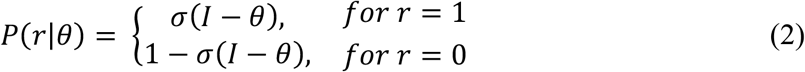

This modeling choice captures the biological variability of MEP responses near the motor threshold, with the probability of observing a motor response increasing sigmoidally as the stimulation intensity *I*surpasses the threshold *θ* [23]. When no response is observed (*r* = 0), the likelihood is the complement of the sigmoid probability. When *I*→ *θ*, there is a 50% chance of eliciting a response.

### Posterior Update

After a binary response, the posterior distribution is updated using Bayes’ rule:

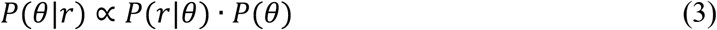

Where *P*(*θ*) is the prior over the threshold *θ, P*(*r*|*θ*) is the likelihood of observing response *r* given threshold *θ*, and *P*(*θ*|*r*) is the posterior after the update. The likelihood *P*(*r*|*θ*) is derived from the sigmoid function with added noise to reflect measurement variability. To keep computation efficient, the posterior is dynamically focused on its peak by adjusting the range of threshold values under consideration. Following sequential Bayesian inference, the posterior from each trial serves as the prior for the next and is updated by combining it with the likelihood derived from the newly observed MEP response. The next stimulation intensity is selected as the posterior mean, representing the most probable motor threshold based on all accumulated evidence. This adaptive process enables BUDAPEST to efficiently converge on the underlying threshold while minimizing unnecessary stimulations [20,24,25].

### Uncertainty Estimation

In medical AI applications, such as diagnostics and medical image reconstructions, uncertainty estimation is used to assess prediction confidence and determine whether results are sufficiently reliable when ground truth is unavailable [26,27]. This is especially relevant where model uncertainty can highlight low-confidence regions and support clinical interpretation. Analogously, BUDAPEST incorporates an uncertainty-based stopping criterion to guide MT estimation, terminating with confidence. At each trial, uncertainty is quantified as the standard deviation of the posterior distribution, reflecting the algorithm’s convergence of the estimate. In this study, we test multiple stopping criteria with virtual subjects and predefine the stopping criteria at 2% uncertainty for human experiments, corresponding to an expected MT accuracy within approximatley ±2% of the underlying threshold.

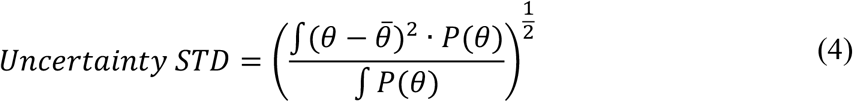

Where 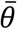 is the posterior mean.

### Trial-by-Trial Refinement of the MT Estimate

The iterative behavior of the BUDAPEST algorithm is illustrated in Figure 2, which shows the evolution of the posterior MT distribution during a representative estimation run. The posterior is initially broad, reflecting high uncertainty, and progressively narrows as additional TMS pulses are incorporated, indicating increasing confidence in the estimate. By Trial 10, the distribution is sharply peaked, signifying convergence to the final MT. Once the posterior standard deviation falls below the user-defined uncertainty threshold, the algorithm alerts the user through the GUI (with an audible notification indicating convergence). This visualization demonstrates BUDAPEST’s ability to efficiently reduce uncertainty and converge on an accurate MT estimate in a small number of trials.

**Figure 2:**
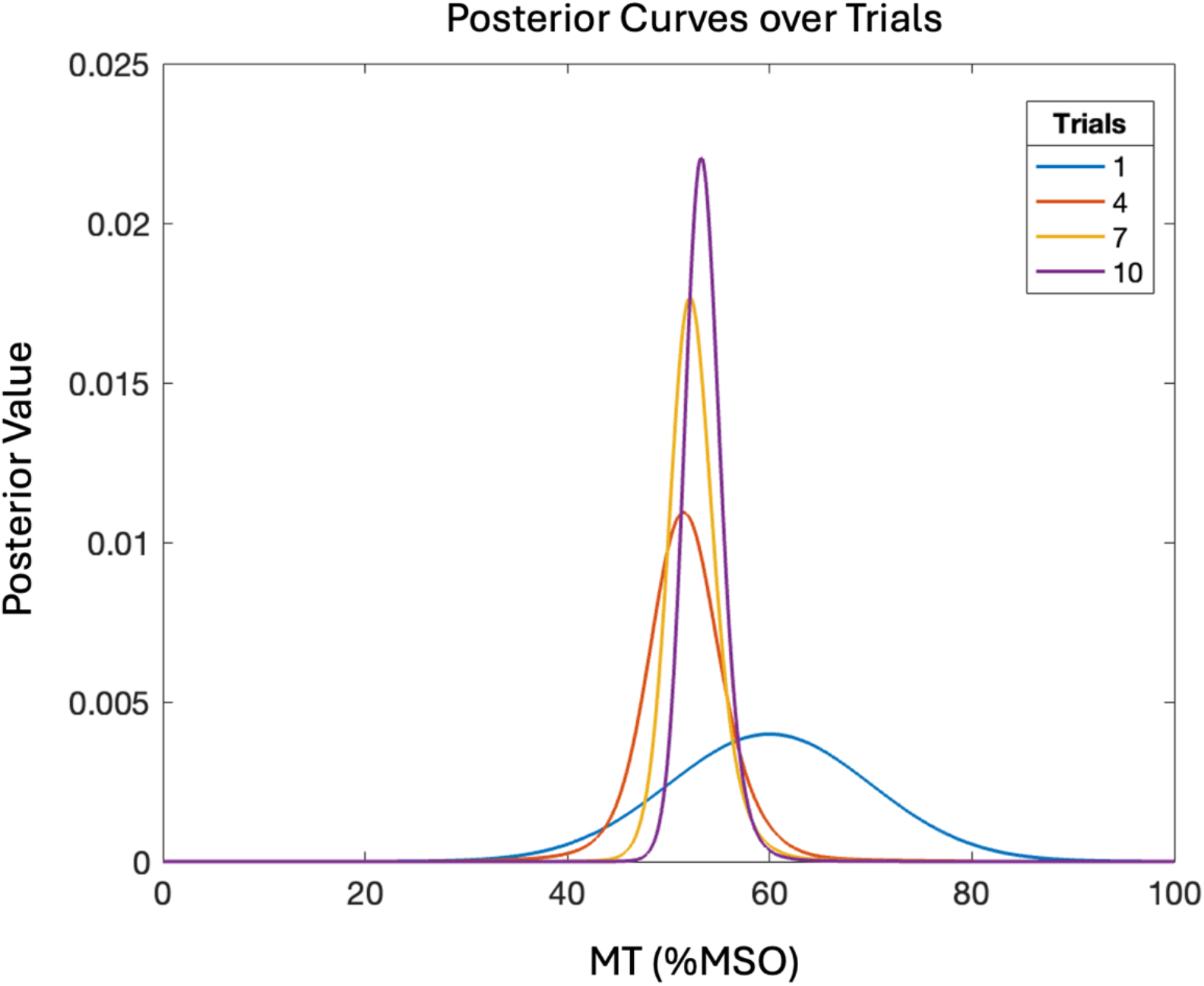
Evolution of the Posterior Distribution. The plot shows the posterior probability distribution of the MT at different stages of the BUDAPEST estimation process (Trials 1, 4, 7, and 10). With each successive trial, the distribution becomes narrower and more peaked, visually representing the reduction in uncertainty as the algorithm converges on the final MT estimate.

### Experimental Protocol

#### Virtual Subject Simulations

The performance of the BUDAPEST algorithm was first evaluated through simulations using virtual subjects to assess its accuracy and convergence speed under varying stopping criteria. Following the approach of Koponen et al., who generated 100 virtual subjects with true MT values ranging from 44% to 84% MSO to evaluate the PEST algorithm [19], we generated 100 virtual subjects and simulated 100 independent runs per subject, resulting in a total of 10,000 virtual experiments. Initial stimulation intensities were randomly selected either 10–15% MSO below or 10-15% MSO above the true MT to evaluate robustness to biased starting intensities, a known limitation of conventional threshold estimation methods. Simulations were conducted using three uncertainty-based stopping criteria, defined as posterior standard deviations of 1%, 2%, or 5% MSO. These criteria allow users to specify a trade-off between estimation accuracy and procedure duration based on experimental requirements.

#### Experimental Setup

To determine the MT and evaluate its spatial variability across the motor cortex, we conducted a series of TMS experiments using both manual hotspot identification and the BUDAPEST algorithm. Prior to the TMS session, structural MRI data were acquired using a 3T Siemens Skyra scanner. High-resolution T1-weighted images were obtained using a MEMPRAGE sequence with the following parameters: repetition time (TR) = 2530 ms, echo time (TE) = 1.76 ms, flip angle = 7°, field of view (FOV) = 256 × 256 mm^2^, and isotropic voxel resolution of 1 × 1 × 1 mm^3^. These MRI data were used for neuronavigation with the Localite TMS Navigator system (Localite, Bonn, Germany), enabling accurate spatial guidance of TMS coil positioning throughout the experiment. TMS was delivered using a Cool-B35 coil (MagVenture, Farum, Denmark; outer diameter 2×46 mm) connected to a MagPro X100 stimulator (MagVenture). Surface electromyography (EMG) recordings were obtained from the first dorsal interosseous (FDI) muscle of the dominant hand using TMS-compatible Ag/AgCl C-shaped electrodes (Easycap GmbH, Wörthsee, Germany) in a bipolar belly–tendon configuration. The ground electrode was placed over the ulnar styloid process. Before electrode placement, the skin was prepared with abrasive gel to maintain impedance below 10 kΩ. EMG signals were recorded, amplified (1000×), band-pass filtered between 20–1000 Hz, and digitized at 20 kHz using the Bittium NeurOne system (Oulu, Finland).

The BUDAPEST algorithm and its accompanying GUI were developed in MATLAB R2024b and operate on standard computing hardware. The system interfaces with the MagVenture TMS stimulator via a TERA USB-C to RS-232 serial adapter, enabling automated control of stimulation parameters directly from the GUI. The GUI was integrated with MAGIC, an open-source MATLAB toolbox for external control of TMS devices, allowing seamless execution of amplitude adjustments and pulse triggering through programmable buttons [28].

#### Resting and Active Motor Threshold Estimation

MT was measured under both resting and active muscle conditions. For rMT, participants were instructed to completely relax their hand and forearm muscles during stimulation. No voluntary contraction was present, and EMG signals were monitored to confirm a low baseline, ensuring minimal muscle activity. MEPs were defined as the peak-to-peak amplitude of EMG responses occurring within 20–50 ms after each TMS pulse [29]. A rMT response was classified as present if the peak-to-peak MEP amplitude exceeded 50 μV. In contrast, aMT was assessed while the subject gently held a small coin between their fingers, maintaining a consistent grip that produced a baseline EMG signal of approximately 100 μV. This mild voluntary contraction ensured continuous activation of the target muscle. For aMT, an MEP response was classified as present only if the peak-to-peak amplitude exceeded 200 μV [30,31]. This distinction accounts for the increased excitability of the motor system during voluntary contraction. The traditional “5-out-of-10” method [12] was treated as the ground truth for evaluating the accuracy and convergence of the BUDAPEST algorithm.

#### Hotspot Identification

Stimulation began at a low intensity to avoid patient discomfort and reduce the likelihood of eliciting MEP responses from an incorrect hotspot location due to excessively high intensity. We applied single TMS pulses at randomly selected cortical locations in the vicinity of the motor hand area, specifically around the precentral gyrus and presumed motor sulcus region. If no EMG response was observed, the intensity was gradually increased in small increments until an MEP response (≥50 μV) was elicited. Once a response was detected, we systematically explored surrounding cortical areas to locate the “tentative hotspot”, the site that produced the highest and most consistent MEPs. For all stimulation sites, the coil orientation was kept at 90 degrees to the central sulcus.

#### Motor Threshold Estimation Using the BUDAPEST Algorithm

The BUDAPEST algorithm was first performed at the tentative hotspot (grid center, as shown in Supplementary Figures S1– S5) to estimate the motor threshold. Binary EMG responses (MEP ≥ 50 μV = 1, otherwise = 0) were used to iteratively update a posterior probability distribution over a range of candidate threshold values.

#### Grid Mapping

After identifying the tentative hotspot, we placed a 3×5 stimulation grid centered on this location, with each grid point spaced 7 mm apart in both anterior-posterior and medial-lateral directions. The TMS coil was kept tangential to the scalp, with the induced E-field direction fixed and perpendicular to the overall orientation of the central sulcus, corresponding to the typical direction that activates corticospinal neurons. Each grid point was stimulated independently using the BUDAPEST algorithm to estimate the motor threshold at that specific location. Any site with stimulation intensity above 150% of the estimated hotspot rMT was deemed out-of-bounds (OOB) and excluded from further analysis to avoid unnecessary subject discomfort (per IRB approved protocol). Both rMT and aMT were recorded for all grid positions.

#### Convergence and Initialization Analysis

To assess the robustness of the BUDAPEST algorithm across different starting intensities, we performed additional runs of the algorithm at the hotspot using starting intensities offset from the true MT by ±10% and ±15%. Each initialization was used as the starting stimulation intensity and convergence accuracy was quantified to evaluate the sensitivity of the algorithm to starting conditions.

## Results

### Accuracy with Simulated Subjects

The BUDAPEST algorithm demonstrated high accuracy and precision in estimating the MT across all uncertainty stopping criteria tested with virtual subjects (Table 1), offering a performance advantage over traditional adaptive procedures such as PEST. BUDAPEST maintained controlled and predictable estimation performance across all stopping criteria. The 2% uncertainty threshold, selected for human experimentation, demonstrated favorable balance between accuracy and robustness, achieving a low mean absolute error (MAE) of 1.93% MSO, 95% error interval of −6.0 to 5.0% MSO, and precision using posterior STD of 2.56% MSO. Worst-case errors under the 2% threshold were well-bounded (±14% MSO), with no tendency toward persistent underestimation as observed in PEST. The more stringent 1% criterion further reduced error (MAE = 1.27% MSO; 95% error interval −4.0 to 3.0% MSO; posterior STD = 1.69% MSO) but at the cost of increased trial count. This performance is comparable to that reported for SAMT in current clinical studies, both in terms of accuracy and the number of trials required for convergence [22]. The 5% threshold produced larger variability (MAE = 3.84% MSO; posterior STD = 4.60% MSO) but still avoided the extreme misestimation behavior seen in PEST.

**Table 1.** Simulation performance of the BUDAPEST algorithm across uncertainty-based stopping criteria. Results summarize the trade-off between estimation accuracy (MAE), precision of the final MT estimate (posterior standard deviation), and convergence efficiency (average number of trials). Data reflect 10,000 simulated threshold estimations across 100 virtual subjects.

| BUDAPEST Stopping Criteria |  |  |  |
| --- | --- | --- | --- |
| Stopping Criteria<br>STD | Mean Absolute Error | Results STD | Avg Trials |
| 1% | 1.274 | 1.69 | 26.4 |
| 2% | 1.932 | 2.56 | 10.2 |
| 5% | 3.840 | 4.6 | 3.6 |

Convergence speed also varied systematically with the stopping criterion, reflecting a trade-off between efficiency and precision (Table 1, Figure 3). As shown in Figure 3a, a lenient stopping criterion of 5% uncertainty produced the fastest convergence, requiring an average of only 3.6 trials, with most runs terminating within 3–5 pulses. However, this speed was achieved at the cost of reduced accuracy. The strict 1% criterion produced the most stable and accurate estimates but required substantially more sampling, converging in 24–30 trials (mean = 26.4). The 2% criterion offered practical balance between speed and precision, with convergence consistently achieved in 8–12 trials (mean = 10.2), closely aligning with an efficient experimental runtime.

**Figure 3.**
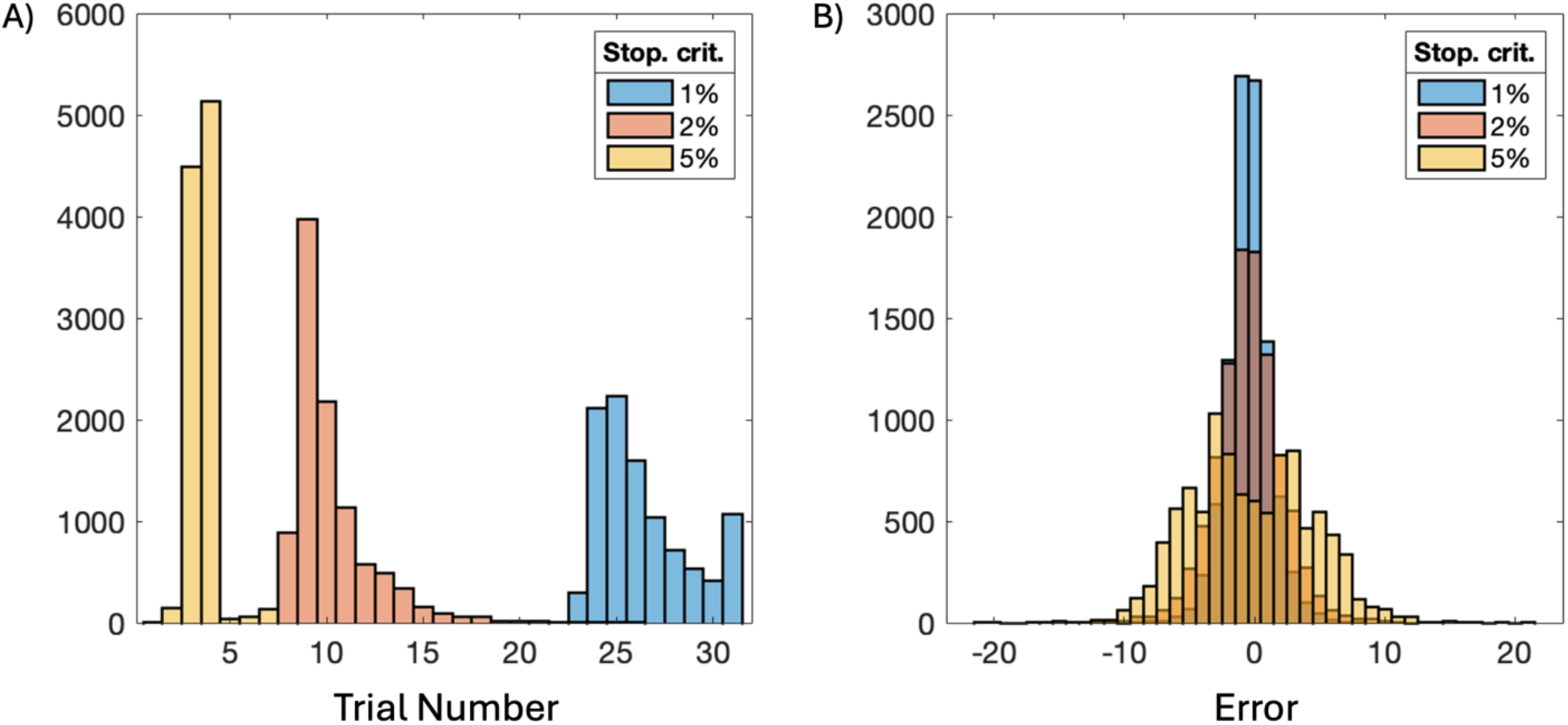
Simulation performance of the BUDAPEST algorithm. (A) Number of trials required for convergence decreases as the uncertainty threshold is relaxed, with the 2% criterion achieving a balance between rapid convergence and stability. (B) Error distributions are centered near zero for all criteria, indicating unbiased estimation, while precision decreases as the stopping threshold becomes more lenient.

Figure 3b further illustrates the error distributions for each stopping criterion. All produced error distributions centered around zero, indicating that BUDAPEST remained unbiased regardless of stopping rule. However, the dispersion of the distributions varied, mirroring the precision metrics reported in Table 1. The 1% criterion produced the narrowest spread of errors, while the 5% criterion resulted in the broadest error distribution. The 2% criterion provided an optimal compromise, maintaining controlled variability while minimizing the number of required stimulation pulses. These results validate the use of a 2% uncertainty threshold as a practical default, enabling rapid MT estimation without compromising accuracy.

Other adaptive Bayesian methods have demonstrated even faster convergence in MT estimation. For example, one study reported that a Bayesian PEST approach using a population-level prior, derived from prior MT measurements within the same laboratory or institution, converged in approximately seven trials, while incorporating subject-specific priors from previous sessions reduced convergence to as few as three trials. In the latter case, the prior primarily served to confirm an already established MT [20]. Although effective, such approaches rely on prior MT data, which may not always be available or generalizable across experimental or clinical settings. In contrast, BUDAPEST converges in approximately 10 trials without requiring prior data, making it well suited for first-time MT estimation and diverse participant populations.

### Experimental Validation in Human Participants

With human participants, the BUDAPEST algorithm was performed using the 2% uncertainty stopping criterion for both rMT and aMT, with the “5-out-of-10” method serving as the ground truth. As shown in Figure 4a, rMT measurements (n = 60 MT estimations) exhibited tightly bounded estimation error, with 50% of estimates falling within ±2% MSO of the experimenter-confirmed MT and all estimates contained within ±4% MSO. AMT measurements (n = 17 MT estimations; error range: −4% to +3% MSO) demonstrated a comparable error distribution, though with slightly greater variability, consistent with increased trial-to-trial variability in MEP responses during voluntary muscle contraction.

**Figure 4.**
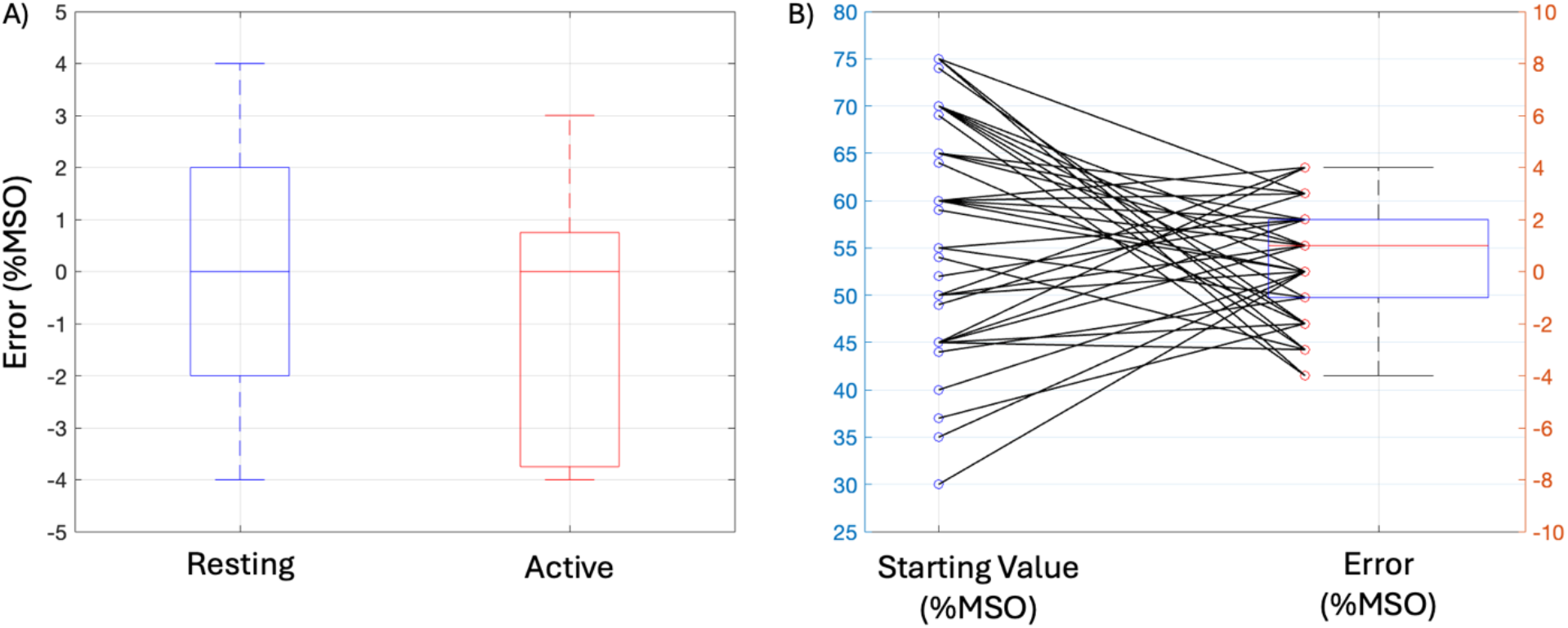
Accuracy and robustness of BUDAPEST in human motor threshold estimation. (A) Estimation error for resting (n = 60) and active (n = 17) MT measurements using the 2% uncertainty criterion. Errors were tightly bounded within ±4% MSO, demonstrating stable accuracy across MT paradigms. (B) Convergence behavior across 50 MT estimation runs with intentionally offset starting intensities. The algorithm consistently converged to accurate MT estimates regardless of whether the initial intensity was above or below the true MT, confirming robustness to initialization error.

To evaluate robustness to poor initialization, we conducted 50 MT estimation runs across five human participants, each intentionally started at a stimulation intensity substantially above or below the true MT to emulate realistic operator error. As illustrated in Figure 4b, BUDAPEST consistently converged to accurate MT estimates regardless of whether the initial value was overestimated or underestimated, demonstrating insensitivity to initial conditions. The crisscrossing convergence trajectories highlight stable directional correction across runs. The overall convergence error exhibited low variability (STD = 2.24% MSO), closely matching simulation results (STD = 2.56% MSO for the 2% criterion), and demonstrated minimal estimation bias (median error = +1% MSO). Furthermore, 50% of MT estimates fell within −1% to +2% MSO of the final accepted MT, and nearly all estimates were contained within a ±4% MSO range, confirming high precision. Collectively, these findings validate BUDAPEST in human experiments and establish it as a rapid, accurate, and initialization-independent MT estimation method, overcoming limitations of conventional approaches that rely on accurate starting intensities.

### Comparison of Resting vs. Active Motor Threshold and Session-to-Session Reliability

The relationship between rMT and aMT is shown in Figure 5a. Across all 3×5 (n=15) grid stimulation sites and 5 participants, aMT values were generally lower than rMT, consistent with increased corticospinal excitability during voluntary muscle contraction and literature [31]. This relationship is reflected by the majority of data points falling below the line of identity (y = x), indicating that less stimulation intensity is required to evoke a motor response in the active state. A significant positive linear relationship was observed between rMT and aMT (R^2^ = 0.55, Pearson r = 0.74), with the regression model aMT = 0.63 × rMT + 19.87 indicating that individuals with higher resting thresholds also tend to exhibit proportionally higher active thresholds. These findings are neurophysiologically consistent [31] and further confirm that the BUDAPEST algorithm produces valid MT estimates across task conditions.

**Figure 5.**
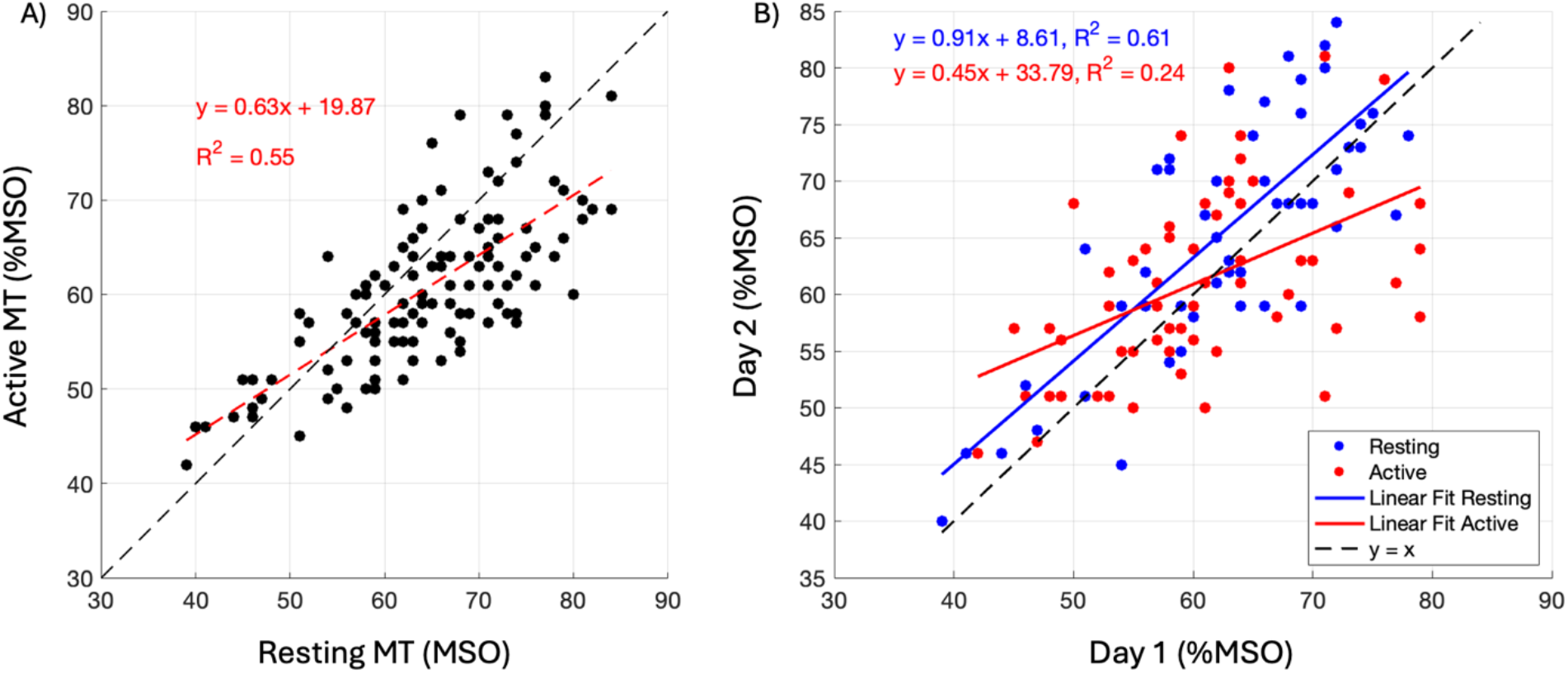
Comparison of resting vs. active motor threshold and session-to-session reliability. (A) Relationship between rMT and aMT across all stimulation sites. AMT values were consistently lower than rMT, as expected due to increased corticospinal excitability during voluntary contraction. A significant positive correlation (R^2^ = 0.55, r = 0.74) indicates that individuals with higher rMT tend to also have higher aMT. (B) Test–retest reliability of MT across two experimental sessions. RMT displayed stronger day-to-day reliability (R^2^ = 0.61, r = 0.78) versus aMT (R^2^ = 0.24, r = 0.49), reflecting greater physiological variability under active muscle contractions.

It is important to note that rMT and aMTs are defined using different electrophysiological criteria. RMT was defined as the lowest stimulation intensity producing MEPs ≥50 µV (peak-to-peak), whereas aMT required MEPs ≥200 µV due to the increased background EMG associated with maintaining a light voluntary contraction. This higher threshold ensures visual detection of MEPs in the presence of ongoing muscle activity. Although these definitions differ, the relationship between rMT and aMT remained systematic and consistent, further supporting the robustness of BUDAPEST-derived estimates across varying physiological conditions.

To evaluate test–retest reliability, rMT and aMTs were measured again on a separate day using identical procedures (Figure 5b). RMT demonstrated strong session-to-session reliability (R^2^ = 0.61, Pearson r = 0.78), with data closely aligned with the identity line and a regression slope near unity (y = 0.91x + 8.61). In contrast, aMT exhibited considerably lower reliability across days (R^2^ = 0.24, Pearson r = 0.49), with greater between-session variability. Due to greater degrees of freedom and additional sources of variability, such as fluctuations in pre-activation force and motor unit recruitment, aMT measurements are more difficult to reproduce consistently across sessions. The reduced reliability of aMT has been reported in prior literature, suggesting that rMT serves as a more stable parameter for standardized and therapeutic applications [31]. Importantly, this variability reflects physiological and experimental instability rather than algorithmic error, as the BUDAPEST algorithm converged reliably under both conditions. Consistent with these findings, rMT emerged as a more stable and reliable metric for longitudinal studies and clinical applications. Notably, this analysis spanned the full 3×5 grid; peripheral sites sample steeper electric-field gradients than the flatter hotspot region, likely reducing reproducibility and correlation. Correlation would therefore be expected to increase further if the analysis were restricted to hotspot-proximal sites.

### Spatial Distribution of Motor Thresholds

MTs were measured over a 3×5 grid centered on the hotspot to evaluate how corticospinal excitability varies spatially across the motor cortex (Supplementary Figures S1–S5). As expected, MT values were lowest at the hotspot (red) and increased with distance from this central location, forming a clear spatial excitability gradient. This pattern was consistent across subjects and supports the physiological interpretation that the hotspot represents the region of strongest corticospinal connectivity, with stimulation efficiency decreasing at peripheral sites due to E-field decay and cortical somatotopic organization. In addition, the OOB sites were also consistently located at the periphery of the map, further demonstrating the monotonic decrease in estimated excitability with increasing distance from the hot spot.

Importantly, within each recording session, the hotspot location was consistent across both resting and active conditions, demonstrating the spatial reliability of the BUDAPEST algorithm for identifying the optimal stimulation site. Across sessions, small shifts in hotspot location were observed in some subjects, typically moving to an adjacent grid position (7 mm spacing). These minor spatial differences fall within normal repositioning variability expected in repeated TMS sessions [32] and are likely due to small changes in head registration and/or coil calibration, rather than algorithmic error. It is interesting to note that the “tentative hotspot” or center of the grid that was determined by initial search did not always correspond to the lowest rMT, suggesting that a systematic approach might be preferable in cases where maximal precision is needed.

## Conclusion and Future Directions

This work introduced the BUDAPEST algorithm, a Bayesian adaptive framework for rapid and reliable MT estimation in TMS. We established its methodological foundation and validated its performance through both simulations and human experiments. BUDAPEST demonstrated high estimation accuracy, with experimental error consistently within ±4% MSO and simulation MAE of 1.9% MSO under the 2% uncertainty criterion. The algorithm also achieved rapid convergence, reaching stable MT estimates in approximately 10 stimulation pulses while maintaining robustness to inaccurate starting intensities, overcoming a major limitation of conventional methods such as PEST.

To enable practical use, BUDAPEST was implemented in an intuitive MATLAB-based GUI that allows experimenters to set user-defined uncertainty thresholds, providing flexible control over the balance between precision and procedure duration. By significantly reducing the number of stimulation pulses required, BUDAPEST decreases overall procedure time and improves participant comfort, a critical consideration in both clinical and high-throughput research environments. Importantly, when operated with a more lenient uncertainty criterion (e.g., 5%), BUDAPEST can converge in as few as 3–4 pulses, providing a rapid, data-driven initialization point that can subsequently be refined using conventional procedures such as the 5-out-of-10 method. This hybrid approach has the potential to substantially reduce the total number of pulses required while preserving the precision of standard MT determination. Faster and more efficient MT estimation may therefore streamline clinical workflows and support timely therapeutic TMS delivery without compromising dosing accuracy.

Finally, we demonstrated the experimental utility of BUDAPEST by comparing rMT and aMT measurements and evaluating session-to-session reliability in human participants. The algorithm produced consistent and unbiased MT estimates, with strong test–retest reliability for rMT. AMT showed greater variability across sessions, reflecting physiological variability rather than algorithmic limitations. These results confirm that BUDAPEST is a robust, accurate, and efficient MT estimation method suitable for both research applications and clinical translation.

The development of a rapid and reliable thresholding algorithm such as BUDAPEST enables new possibilities for scalable, automated, and high-throughput TMS applications. The ability to estimate the MT in approximately one minute per cortical site (e.g., ~10 pulses with a 5-second inter-stimulus interval) makes systematic motor mapping not only feasible but practical for routine experimental and clinical protocols.

A major next step is the integration of BUDAPEST into a fully automated closed-loop TMS framework. In its current implementation, a human operator is required to visually inspect EMG signals and classify MEP responses. This dependency can be eliminated by incorporating real-time automated MEP detection, using either amplitude-based decision rules (e.g., peak-to-peak thresholding) or machine learning classifiers trained to detect MEP waveforms with high sensitivity in noisy conditions [33,34]. Automating response detection would make the thresholding process fully objective and reproducible.

Furthermore, combining BUDAPEST with a robotic TMS coil positioning system (e.g., TMS-Cobot platform by Axilum Robotics) would allow automated multi-site motor mapping [35,36]. In this configuration, the robot would iteratively move the coil across a predefined cortical grid while BUDAPEST estimates MT at each location using online EMG feedback. Such a system could generate a 15-site motor map in ~15 minutes, eliminating operator-dependent variability and dramatically increasing mapping throughput.

BUDAPEST’s Bayesian framework also naturally extends to parallel, multi-muscle threshold inference, whereas existing adaptive MT algorithms such as PEST and SAMT have primarily focused on single-muscle estimation. By recording EMG from multiple muscles simultaneously, a single TMS pulse can update independent threshold posteriors in parallel. Selecting stimulation intensities that efficiently probe the relevant parameter space across muscles could therefore enable concurrent estimation of multiple MTs and support multi-muscle motor mapping within the time typically required to map a single muscle.

This next generation of closed-loop, uncertainty-aware stimulation tools has the potential to substantially advance precision neuromodulation and individualized therapeutic dosing. By reducing manual workload, minimizing observer bias, and enabling fully reproducible TMS procedures, such approaches support standardized neurophysiological biomarkers for clinical trials, rehabilitation and neurorecovery monitoring, and longitudinal assessment of cortical excitability in neurological and psychiatric disorders.

## Data Availability

All data produced in the present study are available upon reasonable request to the authors.

## Acknowledgements

Research reported in this publication was supported by the National Institutes of Health under Award Numbers R01MH128421 (DB, EK, AN, NP), P41EB030006 (AN, MD, NP), R01DC020891 (AN, EK), S10OD028668 (AN), K01MH138823 (MD), The content is solely the responsibility of the authors and does not necessarily represent the official views of the National Institutes of Health. This work was supported in part by The Chernowitz Medical Research Foundation (DJE, AN, EK, DB), and The Pennsylvania Department of Health, Commonwealth Universal Research Enhancement (CURE) Program Award SAP#4100094297 (DJE), as well as the Harvard Brain Science Initiative Bipolar Disorder Seed Grant (FV, AN). We would like to thank Yordan Todorov (MagVenture, Georgia, United States) for supporting the integration of BUDAPEST with the MagVenture stimulator systems.

