## Supplemental Figures for "BUDAPEST: A Fast and Reliable Bayesian Algorithm for TMS Threshold Estimation with an Open-Source GUI and Human Validation"

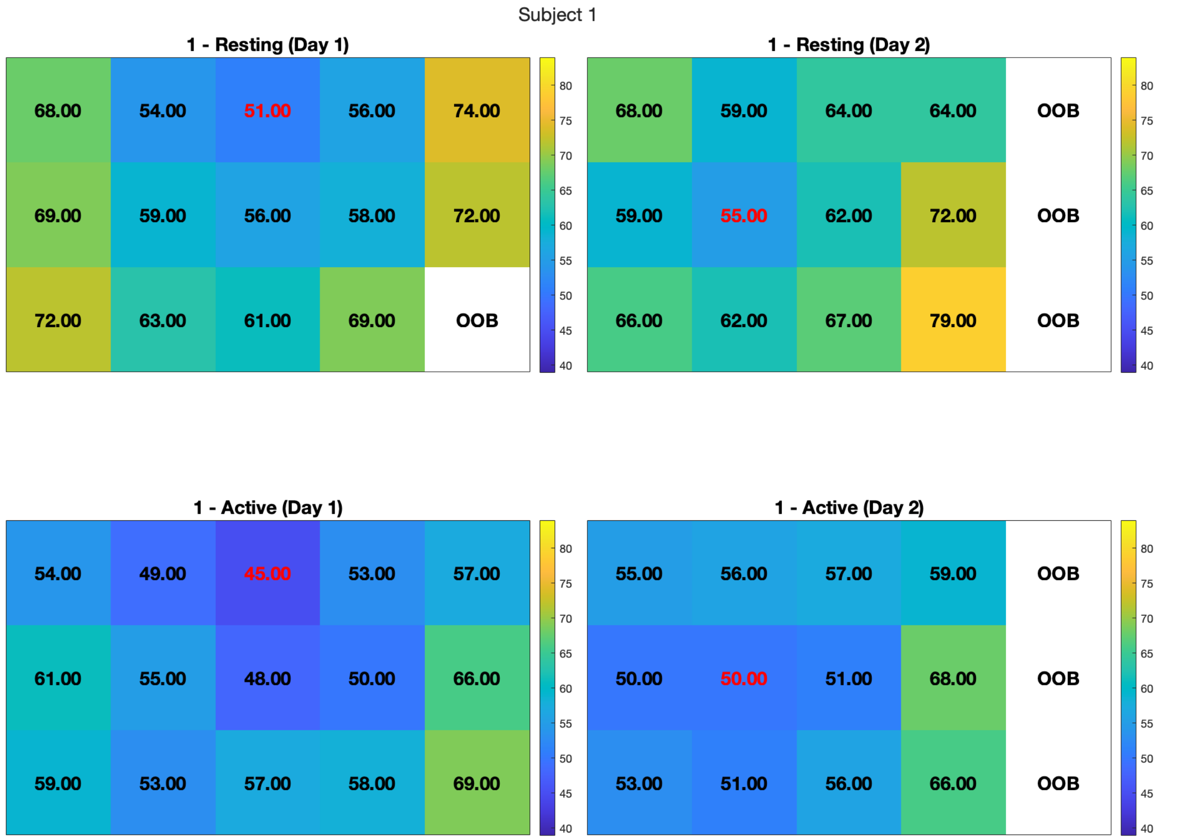


**Supplementary Figure S1. Motor threshold map for Subject 1.**
Motor thresholds (MT) are shown across a 3×5 grid centered on the cortical hotspot (red value). A slight shift in hotspot location is observed between Day 1 and Day 2, with the maximum excitability moving to an adjacent grid position. This shift is within the expected range given that adjacent grid points are spaced 7 mm apart. Such variability is commonly attributed to minor differences in neuronavigation registration, coil positioning, or coil angle across sessions. Despite this positional shift, the overall excitability pattern is stable across days, and MT increases progressively at peripheral grid locations. Out-of-boundary (OOB) values appear only at distant sites where stimulation exceeded the safety limit (>150% of hotspot MT).

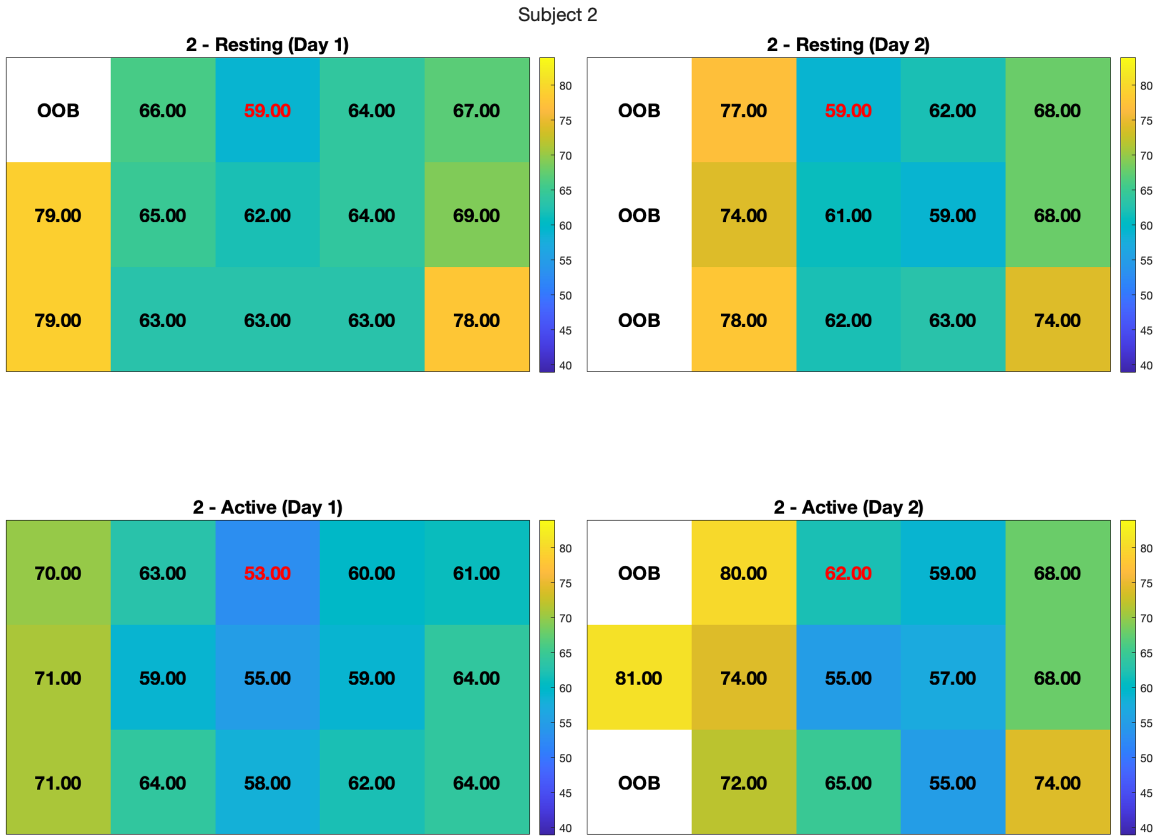


**Supplementary Figure S2. Motor threshold map for Subject 2.**
Spatial distribution of MTs illustrates a clear excitability gradient radiating from the hotspot (red), with thresholds increasing at peripheral grid locations as expected. Day-to-day resting MT values remained stable across sessions, demonstrating strong reliability. However, the active MT at hotspot increased from 53% to 62% MSO between sessions. As shown in the main results, this difference is not due to algorithmic error, BUDAPEST produces accurate MT estimates compared to the standard 5-out-of-10 method, but instead reflects variability inherent to the active MT paradigm. Specifically, active MT depends on maintaining a consistent level of muscle pre-activation, and small changes in voluntary force across sessions introduce variability. This supports our group finding that active MT is less reliable than resting MT due to physiological variability in experimental setup, rather than limitations of the BUDAPEST algorithm.

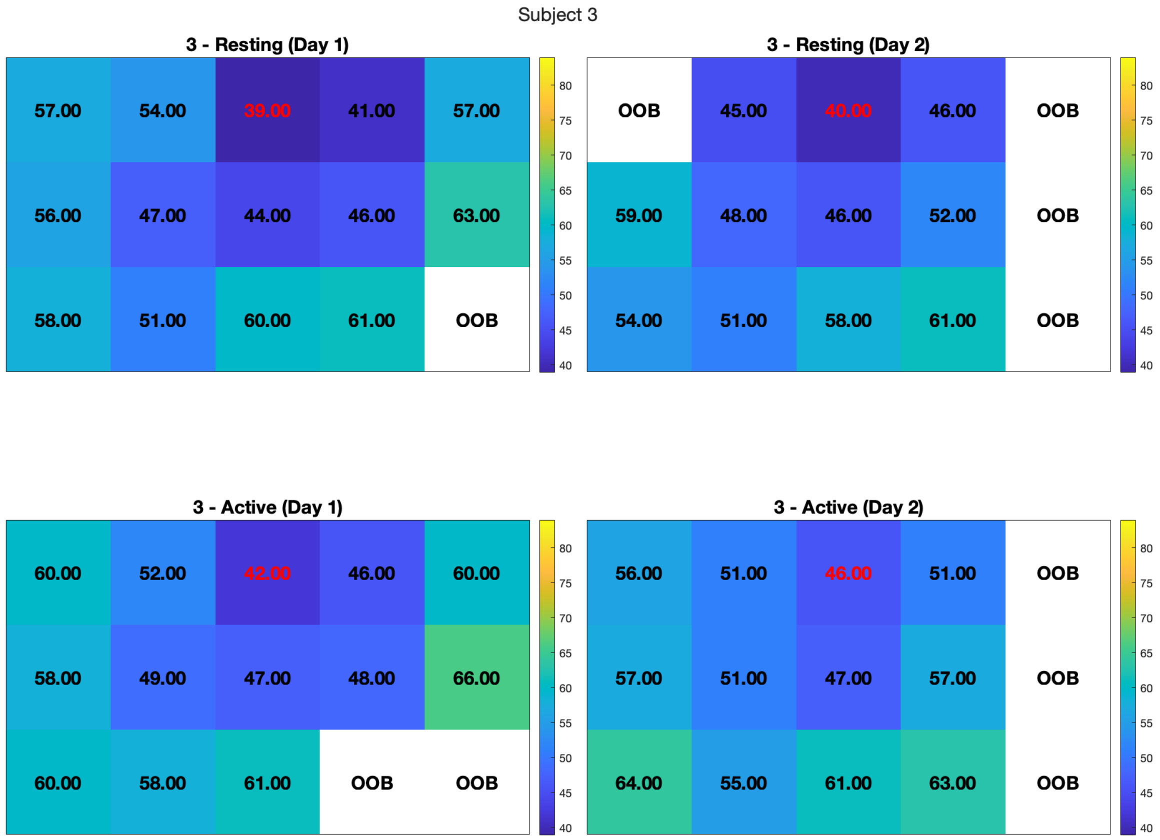


**Supplementary Figure S3. Motor threshold map for Subject 3.**
Both resting and active MT measurements showed strong day-to-day reliability, with the hotspot located in the same grid position across sessions and similar MT values between days. However, in this subject the active MT was sometimes equal to or slightly higher than the resting MT, which deviates from the typical expectation that active MT should be lower due to increased corticospinal excitability. This effect is attributed to the experimental definition of active MT (MEP $\geq$ 200 μV peak-to-peak), which requires the participant to maintain a consistent level of voluntary muscle pre-activation (~100 μV EMG) during each trial. In contrast, resting MT requires a smaller MEP amplitude ($\geq$50 μV) with no background muscle activation, making it inherently more stable and easier to measure. The differing criteria make direct comparisons between resting and active MT challenging, and the variability observed here reflects physiological and experimental inconsistency in active measurements rather than limitations of the BUDAPEST algorithm.

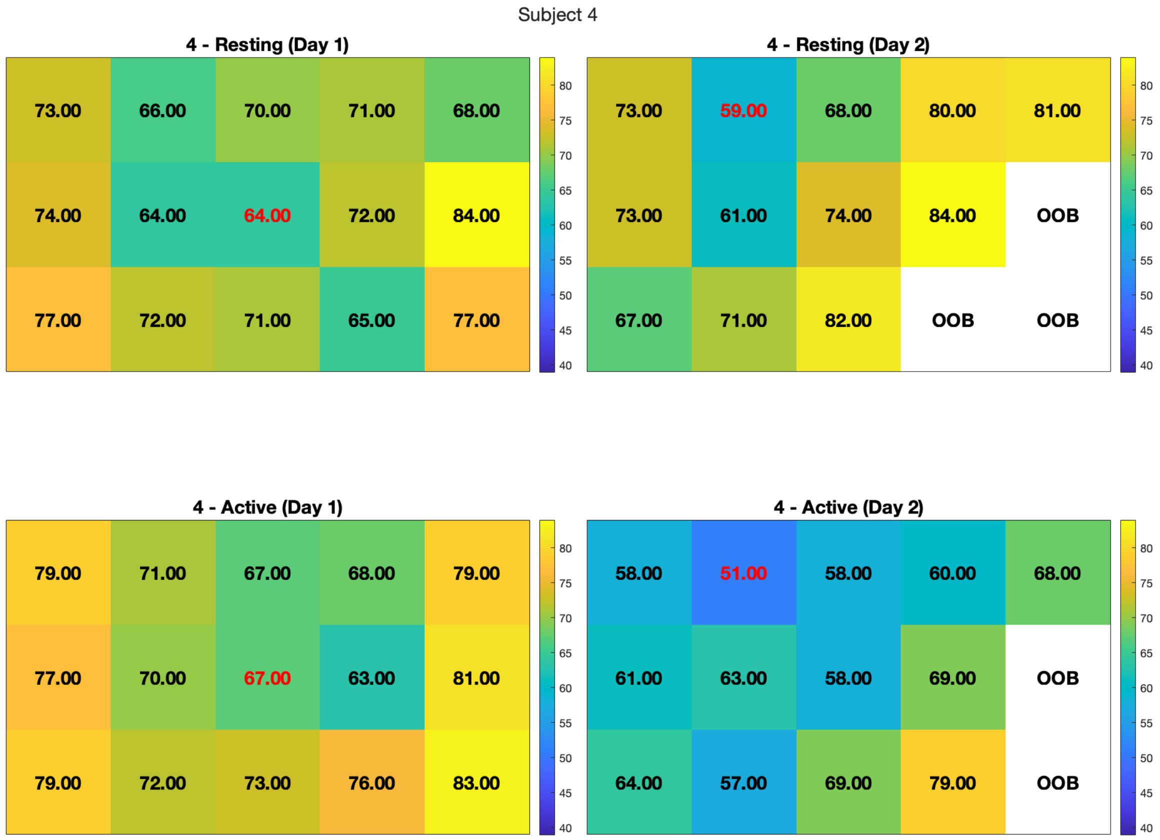


**Supplementary Figure S4. Motor threshold map for Subject 4.**
A slight shift in hotspot location is observed across days, with the site of lowest MT moving to a neighboring grid position. This small spatial shift is within the expected range for repeated TMS sessions and is likely due to normal variability in coil placement, head registration, or small day-to-day differences in head positioning. Resting MT remained relatively stable across sessions and showed a coherent spatial excitability pattern. However, the active MT exhibited a large change between days. As demonstrated in other subjects and in the main analysis, this variability in active MT is not due to variance in the BUDAPEST algorithm but rather reflects the instability of the active measurement itself. This result reinforces that active MT is less reliable than resting MT due to physiological and experimental variability the task demands.

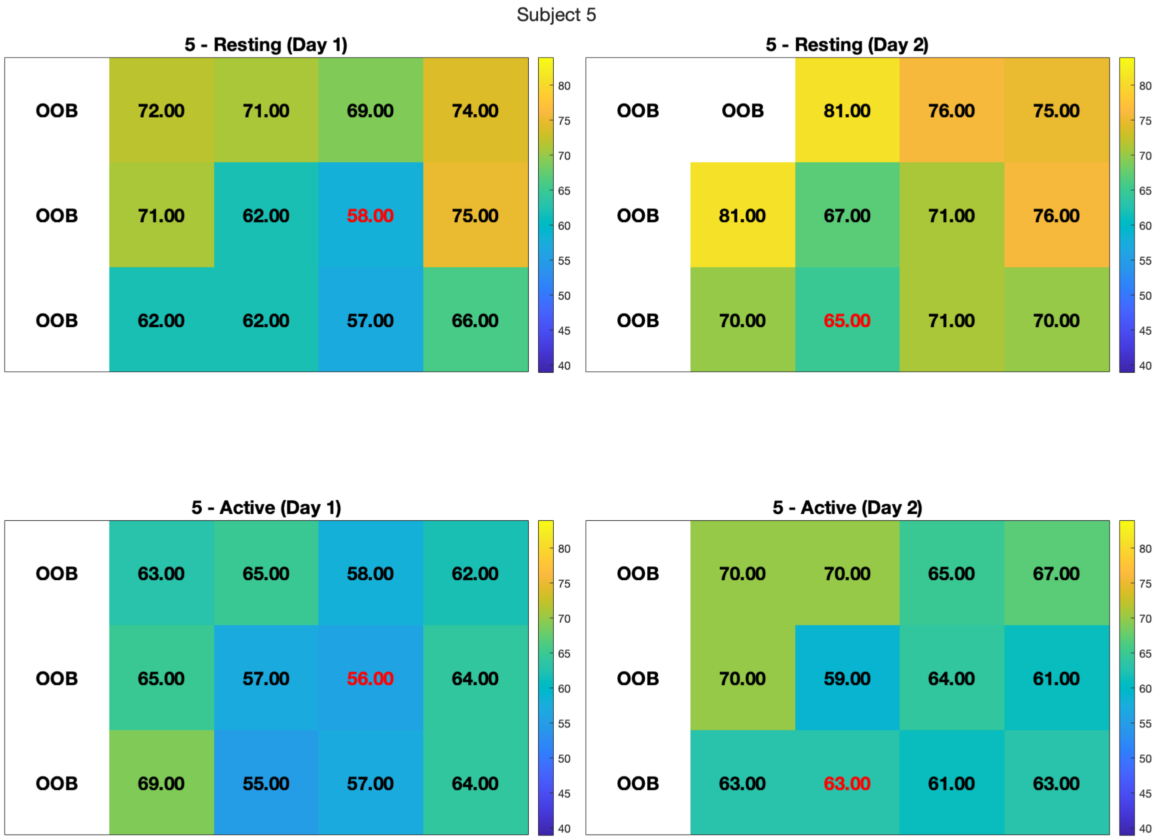


**Supplementary Figure S5. Motor threshold map for Subject 5.**
A small shift in hotspot location is observed between sessions, with the excitability maximum relocating to an adjacent grid position. This is within the expected level of spatial variability across sessions. Resting MT was stable across sessions and showed a consistent excitability gradient centered around the hotspot. In contrast, active MT values changed notably between days. As in previous subjects, this variability reflects the sensitivity of active MT to day-to-day differences in voluntary muscle activation rather than limitations of the BUDAPEST algorithm. These findings further support the conclusion that resting MT provides a more stable neurophysiological measure than active MT for longitudinal studies.
